# Incidence of acute ischemic stroke after COVID-19 or influenza among older adults, findings from PCORnet and HealthVerity, 2022

**DOI:** 10.1101/2024.12.19.24318004

**Authors:** Emilia H. Koumans, Julia Raykin, Tegan K. Boehmer, Sharon Saydah, Jennifer Wiltz, Shikha Garg, Carol E. DeSantis, Thomas W. Carton, Lindsay G. Cowell, Deepika Thacker, Jonathan Arnold, Sonja A. Rasmussen, Samantha J. Smith, Kimberly Barrett, Christine Draper, Fátima Coronado, Elizabeth A. Lundeen, Rebecca C. Woodruff, Jason P. Block

**Affiliations:** Coronavirus and Other Respiratory Viruses Division, National Center for Immunization and Respiratory Diseases, CDC, Atlanta, GA; Office of Public Health Data, Surveillance and Technology, Immediate Office of the Director, CDC, Atlanta, GA; National Center for Chronic Disease Prevention and Health Promotion (NCCDPHP), CDC, Atlanta, GA; Influenza Division, NCIRD, CDC, Atlanta; Louisiana Public Health Institute, New Orleans, Louisiana; Peter O’Donnell Jr. School of Public Health, University of Texas Southwestern Medical Center; Nemours Children’s Health, Wilmington, Delaware; University of Pittsburgh School of Medicine, Department of Medicine, Pittsburgh, Pennsylvania; Johns Hopkins University School of Medicine; Baltimore, Maryland; Harvard Pilgrim Health Care Institute, Harvard Medical School, Boston, Massachusetts; Division for Heart Disease and Stroke Prevention, NCCDPHP, CDC, Atlanta, GA

**Keywords:** ischemic stroke, COVID-19, older adults, complications

## Abstract

**Background:** Although COVID-19 is a known risk factor for thrombotic conditions including embolism, stroke, and myocardial infarction, stroke incidence after implementation of thromboprophylaxis during COVID-19 hospitalization in 2021 and how incidence may differ from influenza is unknown.

**Methods:** PCORnet and HealthVerity (HV) data assets were used to identify patients aged ≥65 years with no prior stroke and COVID-19 or influenza during January 1-December 31, 2022, and AIS from 3 days before to 28 days after COVID-19 or influenza diagnosis. Overall demographic information (age [for HV], sex, race/ethnicity), underlying conditions, level of care, outcomes, and incidence were described and compared between those with early (-3 to 7 days from diagnosis date) and late (8 to 28 days) AIS.

**Results:** Among 245,352 (PC) and 639,396 (HV) patients aged ≥65 years with COVID-19, the incidence of ischemic stroke in the 3 days prior to 7 days after diagnosis of COVID-19 (PC: 962/100,000 and HV: 447/100,000) and influenza (PC: 589/100,000 and HV: 387/100,000) was significantly higher than in the 8 to 28 days after diagnosis (COVID PC: 81/100,000 and HV: 141/100,000)(influenza PC:75/100,000 and HV: 15/100,000)(all P<0.01).

**Conclusions:** Continued risk for AIS with acute COVID-19 and influenza underscore the importance of monitoring and prevention measures for older adults.

## Introduction

Research has consistently found that COVID-19 increases the risk of thrombotic events, such as pulmonary embolism and stroke [1, 2]. Systematic reviews and meta-analyses in 2020 and 2021 estimated that the risk of stroke after COVID-19 ranged from 0.5% to 8.1%, and several studies have reported an increasing risk of acute ischemic stroke (AIS) with increasing COVID-19 severity, such as intensive care unit (ICU) admission [1, 2, 3, 4]. Subsequent meta-analyses and randomized trials have found that COVID-19 and stroke increase the risk for mortality [5], and that therapeutic anticoagulation with heparin improves outcomes among hospitalized patients with COVID-19 [6]. Current treatment guidelines recommend anticoagulation with therapeutic doses of unfractionated heparin or low molecular-weight heparin for acutely ill hospitalized patients with COVID-19 at low risk of bleeding, and prophylactic dosing for all other patients [7, 8]. While there are no thromboprophylaxis recommendations for influenza, excess mortality during influenza season, including from cerebrovascular events, has been recognized since the 19^th^ century [9, 10, 11, 12].

Hypothesized pathophysiologic mechanisms underlying COVID-19-associated stroke include microvascular thromboses, epithelial inflammation, and coagulopathy [1, 2, 3, 4]. These same factors might also be active in influenza [12]. Risk factors for stroke, such as uncontrolled hypertension, diabetes, and cardiovascular disease, increase the risk for hospitalization from COVID-19 and severe COVID-19 outcomes [13]. Consequently, these factors may jointly contribute to both COVID-19 severity and subsequent stroke risk in patients with COVID-19 [1,2,3,4,12]

COVID-19 has been shown to increase the risk of stroke across all age groups, but the risk of stroke is higher in adults aged 65 years and older, and the risk was not well described since the thromboprophylaxis recommendations were released. We examined incidence of AIS after COVID-19 or influenza diagnosis among patients in two large electronic healthcare data assets: electronic health record (EHR) data from The National Patient-Centered Clinical Research Network^*^, PCORnet (PCORnet) and adjudicated insurance claims data from HealthVerity.

Both data assets have information on underlying medical conditions that increase risk for stroke and severe outcomes from COVID-19 or influenza. The study objective was to describe AIS incidence and timing, and characteristics and clinical outcomes of patients aged 65 years or older with COVID-19 or influenza and a diagnosis of AIS, from January 1 through December 31, 2022.

## Methods

### PCORnet

PCORnet provided access to outpatient and inpatient EHR data from 40 sites (each representing one or more health care systems). Participating sites generated aggregate site-level data through a single statistical program package run at participating PCORnet sites; results were returned to the Harvard Pilgrim Health Care Institute, the Coordinating Center for a CDC-funded surveillance project and combined into a single aggregate report with data from all responding sites.

#### Inclusion criteria – PCORnet

The study group included patients aged 65 years or older who were receiving care in PCORnet-affiliated health care systems during January 1-December 31, 2022. Patients with COVID-19 had EHR documentation of a positive SARS-CoV-2 viral test result; an *International Classification of Diseases, Tenth Revision, Clinical Modification* (ICD-10-CM) diagnostic code for COVID-19 (U07.1 or U07.2); or a prescription or administration of a COVID-19 medication (nirmatrelvir/ritonavir or molnupiravir prescription or remdesivir or mAb administration), and no prior COVID-19 diagnosis within 6 months before the index date. Patients with influenza had EHR documentation of a positive influenza test result; an ICD-10-CM diagnostic code for influenza (J09*, J10*, J11*); or prescriptions or administration of influenza medication (oseltamivir or baloxivir). For both conditions, the earliest date of positive test, diagnosis, or therapy was defined as the index date. Patients were excluded if they had any prior ICD-10-CM code for ischemic stroke or transient ischemic attack in any PCORnet records. We examined demographic and clinical information for all COVID-19 and influenza patients from January 1 through December 31, 2022.

### HealthVerity

HealthVerity (https://healthverity.com/) performs privacy-preserving record linkage of patient-level data from over 75 different data partners. We used CDC-licensed HealthVerity adjudicated payor insurance claims data linked to SARS-CoV-2 testing data from commercial laboratories (August 2023 data release). The study period spanned January 1 through December 31, 2022 with a 60-day period through March 1, 2023 for follow-up after acute illness.

#### Inclusion criteria – HealthVerity

The study group included patients aged 65 years and older who were continuously enrolled in a closed payer system from January 1, 2019 until December 31, 2022 and the 60 days following the index date Patients with COVID-19 had a documented COVID-19 ICD-10-CM code (U07.1 or U07.2) or a positive test for SARS-CoV-2 between January 1 through December 31, 2022, and no prior COVID-19 diagnosis within 6 months before the index date. Patients with influenza were defined by a documented ICD-10-CM code for influenza (J09*, J10*, J11*) and no prior influenza diagnosis within 6 months. Patients were excluded if they had any prior ICD-10-CM code for ischemic stroke or transient ischemic attack since January 1, 2019.

Incident AIS was defined as one or more documented ICD-10-CM (I63) codes for ischemic stroke occurring -3 to 28 days from index date. Three days prior because disease onset may occur prior to testing, diagnosis, or treatment for either infection, and up to 28 days because complications may occur after acute illness. Early AIS was defined as occurring -3 to 7 days after the index date, while late AIS was defined as occurring between 8 to 28 days post-index. Supplemental tables present the number and timing of other thrombotic and inflammatory conditions that occurred after COVID-19 and influenza diagnoses with AIS.

### Statistical analyses

The incidence of AIS among patients with COVID-19 or influenza was calculated by dividing the total number of patients with AIS by the total number of patients with COVID-19 or influenza, respectively. Incidence rates per 100,000 people are presented in the text and percents in the Tables and Figure. 95% CI were calculated based on a standard t-distribution. In addition to overall AIS incidence, we further stratified incidence by sex (male and female); age group (for HealthVerity only: 65-74 years, 75-84 years, and ≥85 years); race and ethnicity (non-Hispanic Asian, non-Hispanic Black, non-Hispanic multiracial, non-Hispanic Other, non-Hispanic White, Hispanic, and unknown); illness severity within the three days prior through the 28^th^ day after the index date indicated by an outpatient visit, emergency department visit, hospitalization, or ICU admission (non-exclusive categories). Estimates were additionally stratified by 7 underlying conditions documented at least twice in the previous 3 years: hypertension, type 1 or 2 diabetes mellitus, hyperlipidemia, history of coronary artery disease, obesity, and smoking status (ICD-10-CM codes in supplemental appendix). Outcomes included discharge to hospice care, nursing facility, with home oxygen, or death within 90 days (for PCORnet data)(non-exclusive categories). We compared the proportion of patients with COVID-19 or influenza experiencing AIS in the early period vs. the late period using Pearson’s chi-squared tests. P-values <0.05 were considered statistically significant. This activity was reviewed by the Centers for Disease Control and Prevention (CDC), deemed not research, and was conducted consistent with applicable federal law and CDC policy, US Department of Health and Human Services. Code of Federal Regulations (See e.g., 45 C.F.R. part 46.102(l)(2), 21 C.F.R. part 56; 42 U.S.C. §241(d); 5 U.S.C. §552a; 44 U.S.C. §3501 et seq.. Accessed August 1, 2023 https://ecfr/federalregister.gov/. This study followed standard guidelines for the reporting of observational, cross-sectional studies [14]

## Results

### COVID-19 and ischemic stroke

Overall, 245,352 patients in PCORnet and 639,396 patients in HealthVerity met COVID-19 inclusion criteria (Table 1). Among those with COVID-19, 2,559 in PCORnet and 3,757 in HealthVerity had an ICD-10-CM for AIS in the 3 days prior through the 28^th^ day after the COVID-19 index date. The overall incidence of AIS was 1,043 per 100,000 patients (95% Confidence Interval (CI) 1003-1083) in PCORnet and 588 per 100,000 (95% CI 569-618) in HealthVerity. Characteristics of patients with COVID-19 and AIS from PCORnet and HealthVerity are shown in Table 1; 50% were female; 14% (PCORnet) and 13% (HealthVerity) were non-Hispanic Black, 6.4% (PCORnet) and 9.2% (HealthVerity) were Hispanic, and 54% (PCORnet) and 39% (HealthVerity) were non-Hispanic White. Underlying conditions were common, particularly hypertension (40% PCORnet, 93% HealthVerity). Among those with AIS, at least two-thirds were hospitalized, and 10% or more were discharged to hospice care. Other inflammatory conditions recorded among these patients, by days after index date, are provided Supplemental Table 1.

**Table 1.**
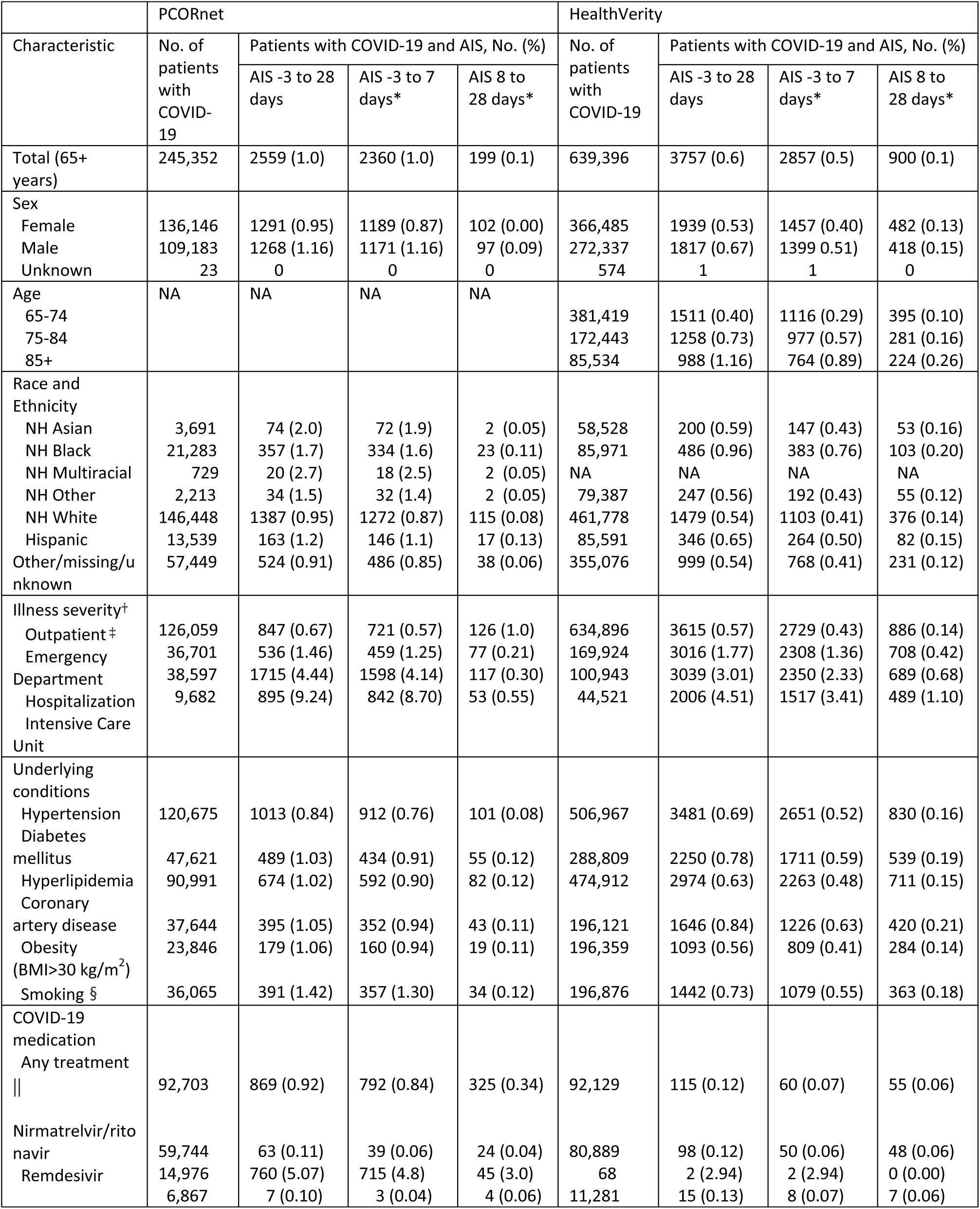
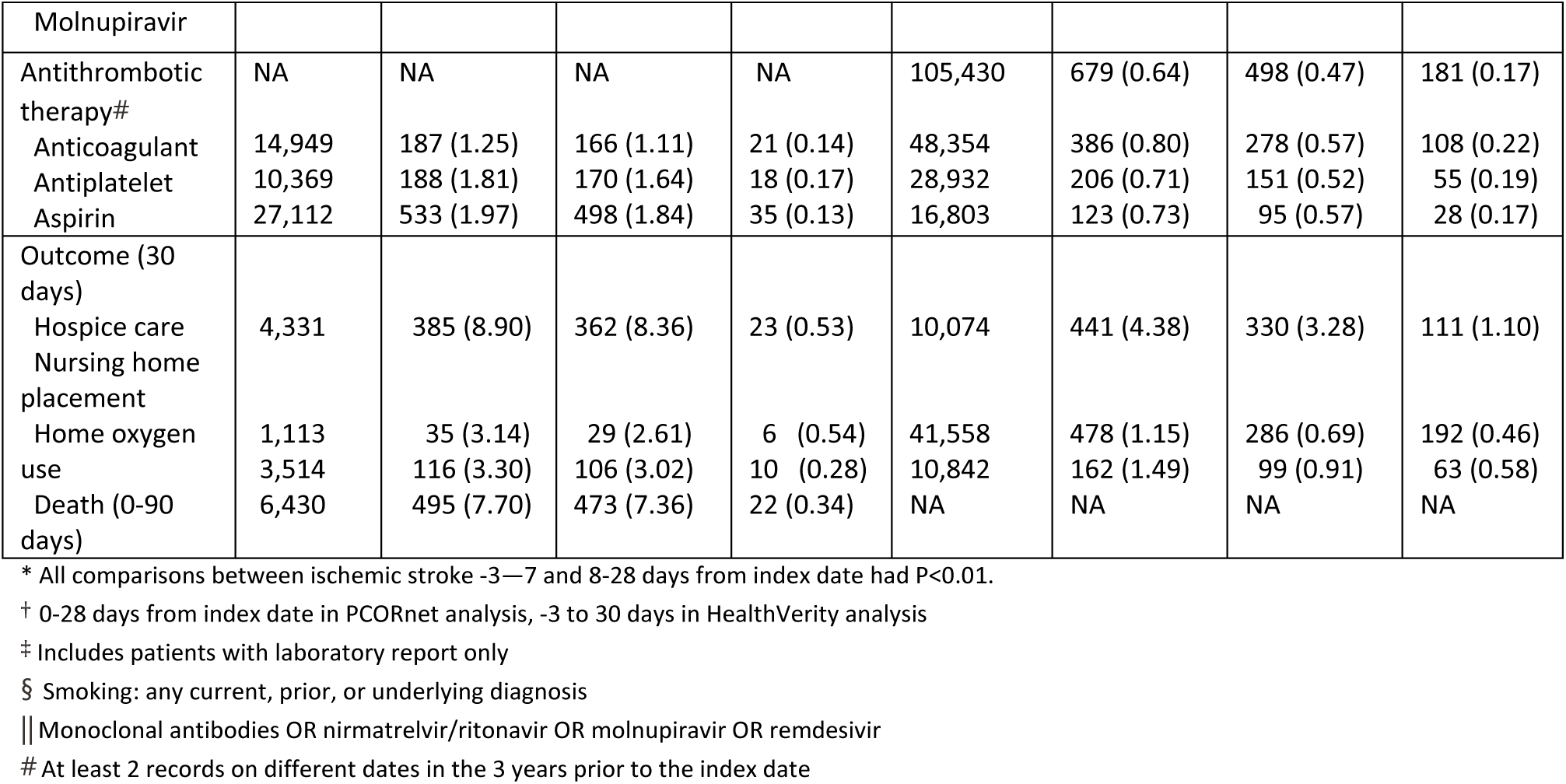
Incidence of acute ischemic stroke among patients aged 65 years and older with COVID-19, by characteristic, in PCORnet (January 1, 2022 through December 31, 2022) and Health Verity (January 1, 2022 through March 1, 2023)

The distribution of age, race and ethnicity, illness severity, underlying conditions, and most outcomes were similar among the groups having early and late stroke (data not shown). Among those with COVID-19 and stroke, 92% (PCORnet) and 76% (HealthVerity) of AIS occurred early (Figure 1). The incidence of early AIS was significantly higher than late AIS in PCORnet (early AIS incidence: 962 per 100,000, 95% CI: 923-101) vs. late AIS incidence: 81 per 100,000, 95% CI: 70-92) and in HealthVerity (early AIS incidence: 447 per 100,000, 95% CI: 430-473 vs. late AIS incidence: 141 per 100,000, 95% CI: 132-156). The incidence of early AIS was significantly higher than the incidence of late AIS among all subgroups examined (Table 1).

**Figure 1.**
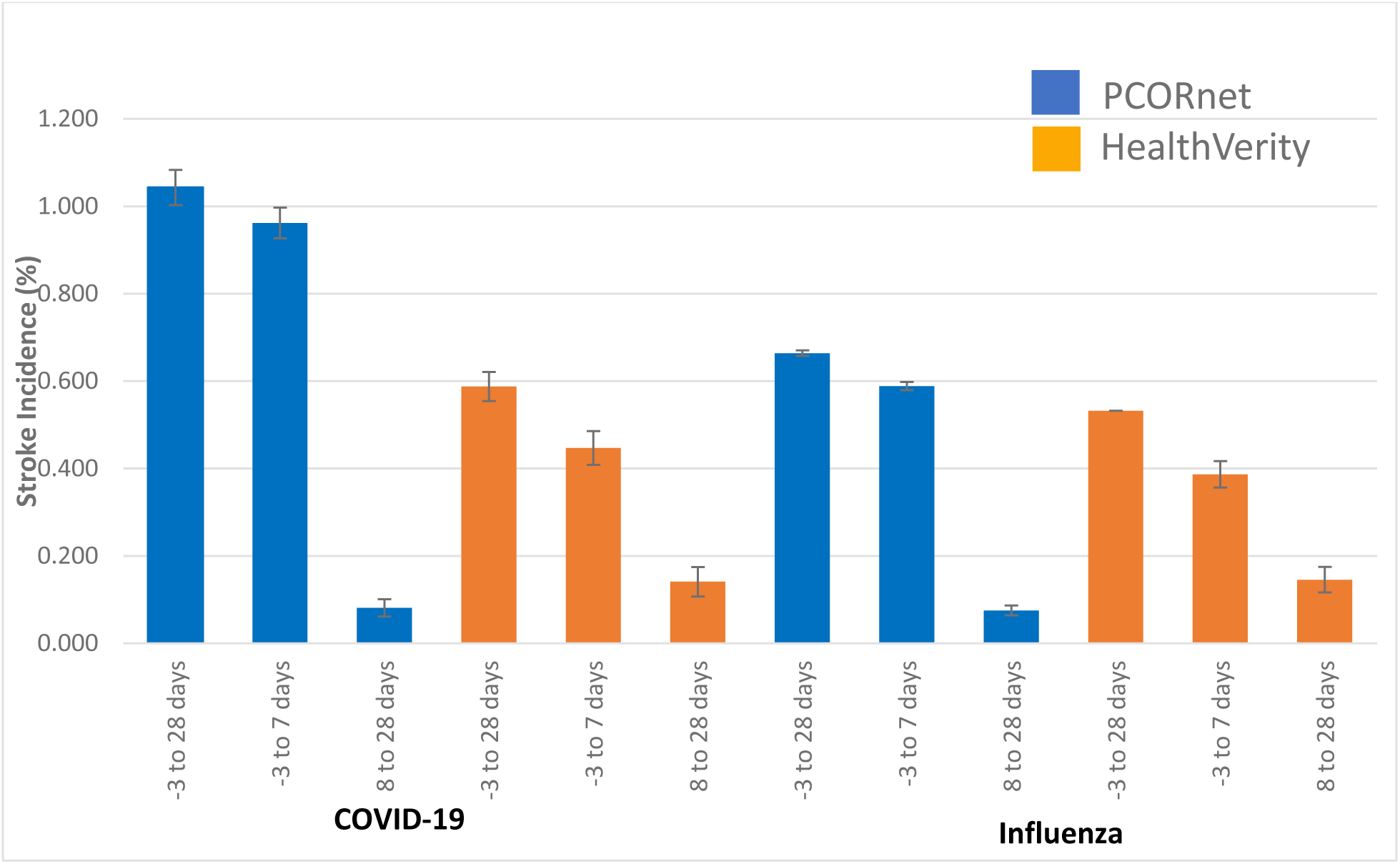
Risk of acute ischemic stroke after COVID-19 and influenza among adults aged 65 years and older, in PCORnet (January 1, 2022, through December 31, 2022) and HealthVerity (January 1, 2022, through March 1, 2023)

### Influenza and ischemic stroke

A total of 14,609 and 59,736 patients met influenza inclusion criteria in PCORnet and HealthVerity, respectively (Table 2). Among those with influenza, 97 in PCORnet and 318 in HealthVerity had an ICD-10-CM code for AIS in the 3 days prior through the 28^th^ day after the index influenza date. The overall incidence of AIS was 664 per 100,000 (95% CI 532-796) in PCORnet and 532 per 100,000 (95% CI 474-591) in HealthVerity. Characteristics of the patients with influenza and AIS from PCORnet and HealthVerity are shown in Table 2; 44% (PCORnet) and 52% (HealthVerity) were female, 22% (PCORnet) and 13% (HealthVerity) were Black, 4% (PCORnet) and 11% (HealthVerity) were Hispanic, and 52% (PCORnet) and 40% (HealthVerity) were White. Underlying conditions were common, particularly hypertension (40%, PCORnet, 94%, HealthVerity). Among those with AIS, at least two-thirds were hospitalized, and 10% or more were discharged to hospice. Other inflammatory conditions recorded among these patients, by days after index date, are provided Supplemental Table 1.

**Table 2.**
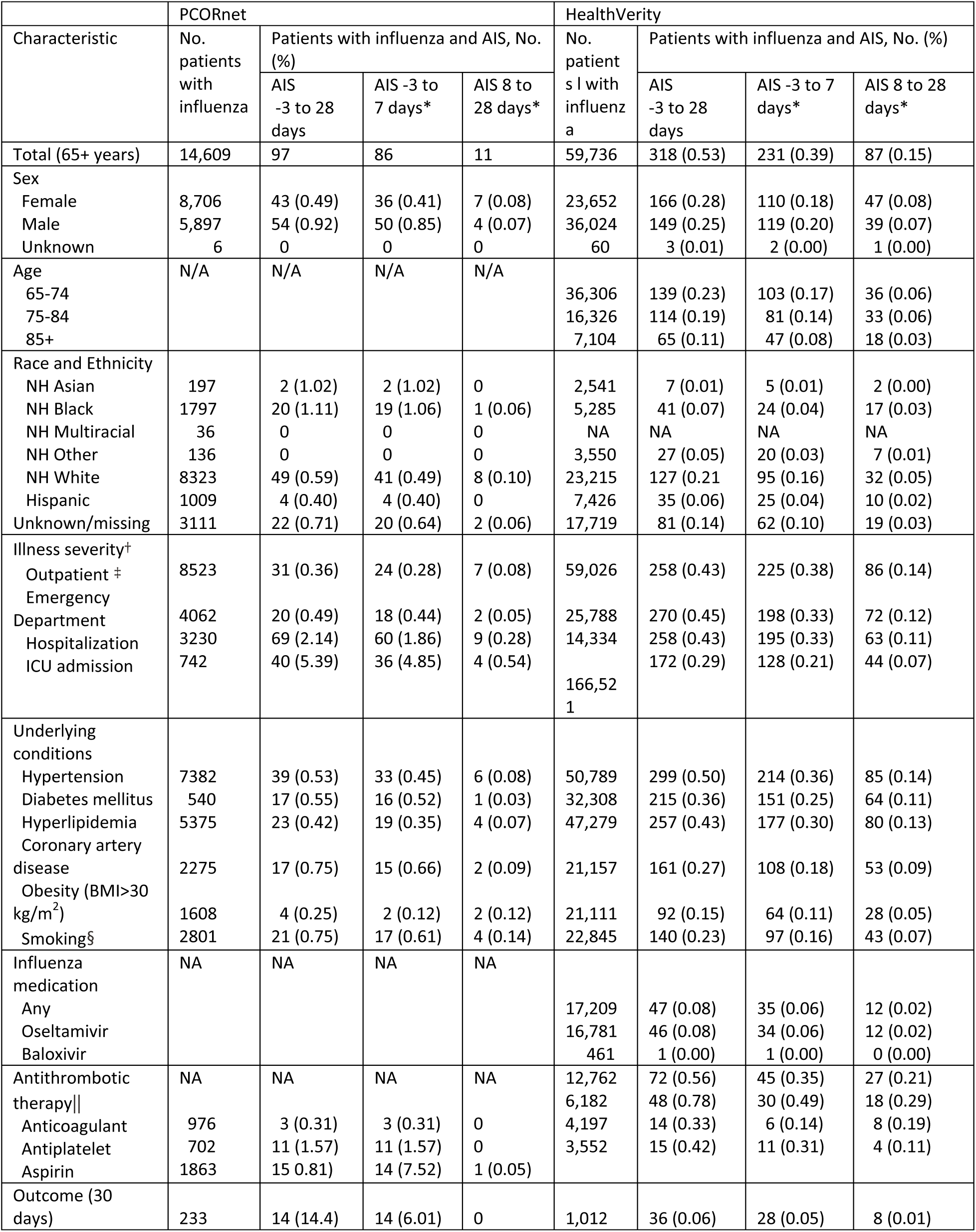
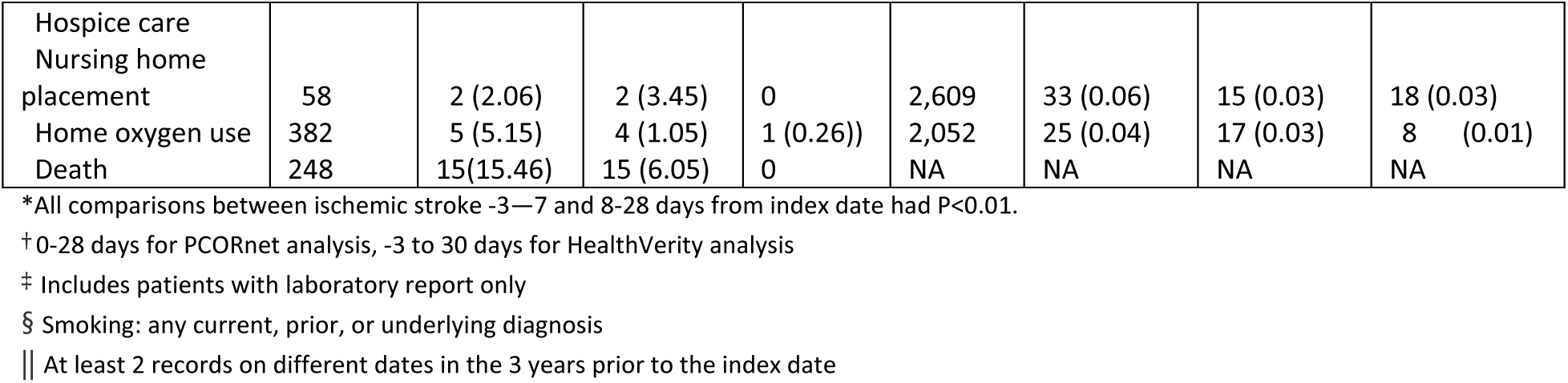
Incidence of acute ischemic stroke among patients aged 65 years and older with influenza, by characteristic, in PCORnet (January 1 through December 31, 2022) and Health Verity (January 1, through December 31, 2022 with follow up through March 1, 2023)

| Characteristic | PCORnet |  |  |  | HealthVerity |  |  |  |
| --- | --- | --- | --- | --- | --- | --- | --- | --- |
|  | No. patients with influenza | Patients with influenza and AIS, No. (%) |  |  | No. patients with influenza | Patients with influenza and AIS, No. (%) |  |  |
|  |  | AIS -3 to 28 days | AIS -3 to 7 days* | AIS 8 to 28 days* |  | AIS -3 to 28 days | AIS -3 to 7 days* | AIS 8 to 28 days* |
| Total (65+ years) | 14,609 | 97 | 86 | 11 | 59,736 | 318 (0.53) | 231 (0.39) | 87 (0.15) |
| Sex |  |  |  |  |  |  |  |  |
| Female | 8,706 | 43 (0.49) | 36 (0.41) | 7 (0.08) | 23,652 | 166 (0.28) | 110 (0.18) | 47 (0.08) |
| Male | 5,897 | 54 (0.92) | 50 (0.85) | 4 (0.07) | 36,024 | 149 (0.25) | 119 (0.20) | 39 (0.07) |
| Unknown | 6 | 0 | 0 | 0 | 60 | 3 (0.01) | 2 (0.00) | 1 (0.00) |
| Age | N/A | N/A | N/A | N/A |  |  |  |  |
| 65-74 |  |  |  |  | 36,306 | 139 (0.23) | 103 (0.17) | 36 (0.06) |
| 75-84 |  |  |  |  | 16,326 | 114 (0.19) | 81 (0.14) | 33 (0.06) |
| 85+ |  |  |  |  | 7,104 | 65 (0.11) | 47 (0.08) | 18 (0.03) |
| Race and Ethnicity |  |  |  |  |  |  |  |  |
| NH Asian | 197 | 2 (1.02) | 2 (1.02) | 0 | 2,541 | 7 (0.01) | 5 (0.01) | 2 (0.00) |
| NH Black | 1797 | 20 (1.11) | 19 (1.06) | 1 (0.06) | 5,285 | 41 (0.07) | 24 (0.04) | 17 (0.03) |
| NH Multiracial | 36 | 0 | 0 | 0 | NA | NA | NA | NA |
| NH Other | 136 | 0 | 0 | 0 | 3,550 | 27 (0.05) | 20 (0.03) | 7 (0.01) |
| NH White | 8323 | 49 (0.59) | 41 (0.49) | 8 (0.10) | 23,215 | 127 (0.21) | 95 (0.16) | 32 (0.05) |
| Hispanic | 1009 | 4 (0.40) | 4 (0.40) | 0 | 7,426 | 35 (0.06) | 25 (0.04) | 10 (0.02) |
| Unknown/missing | 3111 | 22 (0.71) | 20 (0.64) | 2 (0.06) | 17,719 | 81 (0.14) | 62 (0.10) | 19 (0.03) |
| Illness severity† |  |  |  |  |  |  |  |  |
| Outpatient ‡ | 8523 | 31 (0.36) | 24 (0.28) | 7 (0.08) | 59,026 | 258 (0.43) | 225 (0.38) | 86 (0.14) |
| Emergency Department | 4062 | 20 (0.49) | 18 (0.44) | 2 (0.05) | 25,788 | 270 (0.45) | 198 (0.33) | 72 (0.12) |
| Hospitalization | 3230 | 69 (2.14) | 60 (1.86) | 9 (0.28) | 14,334 | 258 (0.43) | 195 (0.33) | 63 (0.11) |
| ICU admission | 742 | 40 (5.39) | 36 (4.85) | 4 (0.54) | 166,521 | 172 (0.29) | 128 (0.21) | 44 (0.07) |
| Underlying conditions |  |  |  |  |  |  |  |  |
| Hypertension | 7382 | 39 (0.53) | 33 (0.45) | 6 (0.08) | 50,789 | 299 (0.50) | 214 (0.36) | 85 (0.14) |
| Diabetes mellitus | 540 | 17 (0.55) | 16 (0.52) | 1 (0.03) | 32,308 | 215 (0.36) | 151 (0.25) | 64 (0.11) |
| Hyperlipidemia | 5375 | 23 (0.42) | 19 (0.35) | 4 (0.07) | 47,279 | 257 (0.43) | 177 (0.30) | 80 (0.13) |
| Coronary artery disease | 2275 | 17 (0.75) | 15 (0.66) | 2 (0.09) | 21,157 | 161 (0.27) | 108 (0.18) | 53 (0.09) |
| Obesity (BMI>30 kg/m<sup>2</sup>) | 1608 | 4 (0.25) | 2 (0.12) | 2 (0.12) | 21,111 | 92 (0.15) | 64 (0.11) | 28 (0.05) |
| Smoking§ | 2801 | 21 (0.75) | 17 (0.61) | 4 (0.14) | 22,845 | 140 (0.23) | 97 (0.16) | 43 (0.07) |
| Influenza medication | NA | NA | NA | NA |  |  |  |  |
| Any |  |  |  |  | 17,209 | 47 (0.08) | 35 (0.06) | 12 (0.02) |
| Oseltamivir |  |  |  |  | 16,781 | 46 (0.08) | 34 (0.06) | 12 (0.02) |
| Baloxivir |  |  |  |  | 461 | 1 (0.00) | 1 (0.00) | 0 (0.00) |
| Antithrombotic therapy\|\| | NA | NA | NA | NA | 12,762 | 72 (0.56) | 45 (0.35) | 27 (0.21) |
| Anticoagulant | 976 | 3 (0.31) | 3 (0.31) | 0 | 6,182 | 48 (0.78) | 30 (0.49) | 18 (0.29) |
| Antiplatelet | 702 | 11 (1.57) | 11 (1.57) | 0 | 4,197 | 14 (0.33) | 6 (0.14) | 8 (0.19) |
| Aspirin | 1863 | 15 (0.81) | 14 (0.75) | 1 (0.05) | 3,552 | 15 (0.42) | 11 (0.31) | 4 (0.11) |
| Outcome (30 days) | 233 | 14 (14.4) | 14 (6.01) | 0 | 1,012 | 36 (0.06) | 28 (0.05) | 8 (0.01) |
| Hospice care |  |  |  |  |  |  |  |  |
| Nursing home |  |  |  |  |  |  |  |  |
| placement | 58 | 2 (2.06) | 2 (3.45) | 0 | 2,609 | 33 (0.06) | 15 (0.03) | 18 (0.03) |
| Home oxygen use | 382 | 5 (5.15) | 4 (1.05) | 1 (0.26)) | 2,052 | 25 (0.04) | 17 (0.03) | 8 (0.01) |
| Death | 248 | 15(15.46) | 15 (6.05) | 0 | NA | NA | NA | NA |

\*All comparisons between ischemic stroke -3—7 and 8-28 days from index date had P<0.01.
† 0-28 days for PCORnet analysis, -3 to 30 days for HealthVerity analysis
‡ Includes patients with laboratory report only
§ Smoking: any current, prior, or underlying diagnosis
|| At least 2 records on different dates in the 3 years prior to the index date

The distribution of age, race and ethnicity, illness severity, underlying conditions, and most outcomes were similar among the groups having early vs late AIS (data not shown). Among those with influenza and stroke, 89% (PCORnet) and 73% (HealthVerity) occurred early (Figure 1). The incidence of early AIS was significantly higher than late AIS in PCORnet (early AIS incidence: 589 per 100,000, 95% CI: 465-713 vs. late AIS incidence: 75 per 100,000, 95% CI:31-120) and HealthVerity (early AIS incidence: 387 per 100,000, 95% CI: 337-436 vs. late AIS incidence: 15 per 100,000, 95% CI: 12-18). The incidence of early AIS was significantly higher than the incidence of late AIS among all subgroups examined (Table 1).

## Discussion

Among patients aged 65 years or older, the incidence of a new AIS was highest during the 3 days prior through the 7^th^ day after the index date for both COVID-19 and influenza. Although AIS incidence was significantly higher during the early period (-3 to 7 days) than during the later period (8 to 28 days) for both illnesses, the magnitude was highest for COVID-19 in PCORnet. Because the PCORnet, but not the HealthVerity sample, included patients who had been treated with a COVID-19-specific medication, more patients with COVID-19 may have been included in PCORnet. The finding of acute stoke occurring the early part of COVID-19 illness align with population-based studies from Israel, Denmark, Sweden, and Scotland conducted in 2020 [15, 16, 17, 18]. In these studies, between 66% and 75% of patients with a stroke were also hospitalized during the acute phase of COVID-19 illness. The overall proportion of patients with AIS following influenza in our study (0.6% to 1.0%) was consistent with findings from pre-COVID-19 pandemic evaluations of non-respiratory diagnoses associated with influenza (0.9%) [17], and from other studies of the risk of stroke with influenza [9-12, 19]; these studies also noted higher stroke incidence during the first 7-10 days of illness, corroborating our finding of heightened stroke risk in the early post-infection period.

This study covered calendar year 2022, which was after the release of recommendations for antithrombotic prophylaxis use for inpatients with COVID-19 in August, 2021 [6]. Unfortunately, we were unable to examine the use of prophylaxis among hospitalized patients to determine its role in the development of AIS. Other studies examining COVID-19-related stroke incidence were predominantly conducted in 2020 and 2021, before the release of thromboprophylaxis recommendations. For example, in a study of hospitalized COVID-19 patients during 2020, the risk of stroke was 4% among 165 patients, and was 5%-6% among those with severe COVID-19 [3]. A meta-analysis using 2020 data reported a pooled incidence of 1.4% for ischemic and hemorrhagic stroke following COVID-19, with a range from 0.5%-8.1% across studies [4]. Our findings fall within the lower range of these studies. Data on stroke incidence from the thromboprophylaxis era or the Omicron era remain sparse. Nevertheless, an increased risk for mortality associated with COVID-19 remains among patients who also have stroke, even with prophylaxis [5].

This study had several limitations. First, these observations are from patients who accessed health care; findings may not generalize to other populations. Second, this study was intended to report surveillance estimates for AIS across the population. We received aggregated data from PCORnet and were unable to adjust for age, other risk factors, or acute therapies, including receipt of thromboprophylaxis, in the incidence of AIS. Third, these data may not have included all occurrences of COVID-19, influenza, and AIS, particularly if patients received care at institutions not captured in the data assets.

In summary, both COVID-19 and influenza were temporally associated with AIS among people aged 65 years and older, with the highest incidence in the 3 days prior to the 7 days after diagnosis. These findings underscore the importance of preventive measures and of close monitoring of older adults with COVID-19 or influenza, especially within the first week of illness when stroke risk is elevated. Vaccinations against COVID-19 and influenza, along with effective management of chronic conditions like diabetes and cardiovascular disease, key contributors to both stroke and severe COVID-19, are essential strategies to reduce thrombotic events. Additionally, raising awareness of the early signs of stroke among patients and healthcare providers is vital for prompt recognition and timely intervention that can significantly improve outcomes.

## Data Availability

Data presented are not available to the public.

## Acknowledgements

Evelyn Twentyman for initiating this project.

No funding was received to assist with the preparation of this manuscript.

## Disclosures

### Non-financial interests

SAR serves on advisory committees for pregnancy registries for medications manufactured by Axsome Therapeutics, Harmony Biosciences, Myovant Sciences, Novo Nordisk, and Pfizer, and receives royalties for an article on influenza and pregnancy from UpToDate.

All authors contributed to the study conception and design. Material preparation, data collection and analysis were performed by Julia Raykin, Carol E. DeSantis, Samantha J. Smith, Kimberly Barrett, Christine Draper, and Emilia Koumans. The first draft of the manuscript was written by Emilia Koumans and all authors commented on previous versions of the manuscript. All authors read and approved the final manuscript.

## Footnotes

* https://pcornet.org/data and https://github.com/PCORnet-DRN-OC/Query-Details/tree/master/Therapeutics%20Query, https://pubmed.ncbi.nlm.nih.gov/33002635/

